# Linking Adolescent Alcohol Use to Adult Behavioral Flexibility: Habitual Action-Selection and Attentional Bias

**DOI:** 10.1101/2025.07.16.25331591

**Authors:** Elena M. Vidrascu, Samantha Dove, Madeline M. Robertson, Kristin N. Meyer, Margaret A. Sheridan, Donita L. Robinson, Charlotte A. Boettiger

**Affiliations:** Department of Psychology & Neuroscience University of North Carolina, Chapel Hill, NC USA 27599; Biomedical Research Imaging Center University of North Carolina, Chapel Hill, NC USA 27599; Bowles Center for Alcohol Studies University of North Carolina Chapel Hill, NC USA 27599; Department of Psychiatry University of North Carolina Chapel Hill, NC USA 27599

**Keywords:** Adolescent alcohol use, behavioral flexibility, attentional bias, habitual action-selection

## Abstract

**Background:** Habitual behavior and attentional bias are distinct cognitive processes that both contribute to inflexible behavior and are commonly observed in addiction. While animal studies provide strong evidence for an association between adolescent alcohol use and impairments in behavioral flexibility in adulthood, such a link in human research has not yet been explored. Moreover, since reduced flexible behavior serves as a risk factor for escalating alcohol intake, continued alcohol consumption use in adulthood may further exacerbate any deficits associated with adolescent drinking.

**Methods:** We used principal component analysis to create composite scores for adolescent and past year alcohol use, based on self-report measures from a healthy adult sample. Group differences in alcohol use were examined in relation to habitual responding (n=71) and attentional bias (n=44) toward non-drug reward cues, using two behavioral flexibility tasks. We used linear regression analyses to explore associations between past year alcohol use and behavioral flexibility outcomes in adults with histories of light versus heavy adolescent alcohol use.

**Results:** Heavy adolescent alcohol use was characterized by earlier drinking onset and higher binge-drinking frequency before age 18. Adults with a history of heavy adolescent alcohol use demonstrated significantly greater habitual responding compared to those with lighter use. Among this group, greater past year alcohol use was also associated with increased difficulty disengaging attention from non-drug reward cues.

**Conclusions:** These results indicate that adolescent and current alcohol use may differentially impact habitual responding and attentional bias towards non-drug reward cues. Notably, this is the first human study to explore both aspects of behavioral inflexibility in relation to different periods of alcohol use within the same adult sample.

## Introduction

Addiction is largely characterized by an impaired ability to flexibly override habit-based actions in favor of goal-directed behavior (McKim et al., 2016). Individuals with alcohol use disorder (AUD) also show greater reward-driven attentional capture by alcohol cues (Anderson, 2021, Bollen et al., 2022), reflecting heightened motivational salience of alcohol-related stimuli (Cofresi et al., 2019). Importantly, AUD is strongly associated with early-life alcohol use, particularly during adolescence (Grant and Dawson, 1998, SAMHSA, 2023). While human studies often fail to account for adolescent alcohol use when investigating deficits in behavioral flexibility among adults with an AUD, this developmental window may be critical in shaping vulnerability. Although evidence for attentional capture is more limited than for habit-based behavior, animal studies clearly link both chronic adult alcohol exposure and adolescent alcohol exposure to deficits in adult behavioral inflexibility (Crews et al., 2019, Gomez et al., 2021, Dannenhoffer et al., 2021, Pochapski et al., 2024, Madayag et al., 2017). Furthermore, reduced flexibility has also been identified as a risk factor for escalating drug or alcohol intake (Istin et al., 2017, Nippert et al., 2024, Shnitko et al., 2020). Thus, it may be that deficits in habitual behavior and reward-driven attention associated with AUD are cognitive risk factors acquired during adolescence, prior to any alcohol use. Alternatively, exposure to alcohol in adolescence and adulthood may independently increase inflexible behavior, with adult exposure moderating the impact of adolescent alcohol use on behavioral outcomes. Probing the origin of cognitive deficits associated with alcohol use in non-addicted populations may help identify early cognitive markers of vulnerability and inform targeted strategies to prevent the progression to alcohol use disorder.

Adolescence is a critical period of neurodevelopment during which behaviors tend to be more impulsive and less governed by deliberate, goal-directed decision-making (Gogtay et al., 2004). It is also a time when heavy alcohol use, including binge drinking, is common and may interfere with the maturation of higher-order regions, particularly those that continue to develop into emerging adulthood (for review see Tetteh-Quarshie and Risher, 2022). The ability to flexibly adapt behavior to changing environmental demands relies on intact prefrontal cortex function. Relatedly, lower prefrontal cortex volume and greater cortical thinning are associated with hazardous alcohol use early in life (Cservenka and Brumback, 2017). Together, these findings suggest that binge drinking during adolescence may disrupt the normative development of cognitive control systems, leading to a bias toward reactive, automatic behavior at the expense of goal-directed action – hallmarks of behavioral inflexibility observed in addiction (Luscher et al., 2020).

Given this link, it may be that similar biases toward habitual behavior are evident in subclinical populations with a history of adolescent alcohol use. To date, little research has examined the possibility that early alcohol exposure in humans is associated with increased habitual behavior. This is in part because traditional neuropsychological tasks designed to assess behavioral or cognitive flexibility challenge participants to switch between rules or tasks, but do not differentiate between goal-directed and habit-based behavior (Hohl and Dolcos, 2024, Uddin, 2021). The Hidden Association Between Images Task (HABIT) task (Boettiger and D’Esposito, 2005), however, measures an individual’s ability to override well-learned, habitual responses in favor of goal-directed behavior under dynamic task demands. Using perseverative errors as an index of inflexibility, we have previously shown that adults with a history of addiction exhibit greater habit formation compared to those without such a history (McKim et al., 2016). Applying this task to adult populations without addiction, but with a history of early alcohol exposure, may help uncover latent effects of adolescent alcohol exposure on behavioral flexibility.

Numerous studies have found greater bias towards alcohol-related and non-drug reward stimuli among individuals with an AUD in reward-conditioning paradigms (Anderson et al., 2011, Anderson, 2021, Garland et al., 2012, Hester and Luijten, 2014). However, this bias towards alcohol cues is often modulated by factors such as abstinence duration and current motivational state, which can complicate efforts looking at long-term effects of adolescent alcohol exposure (Bollen et al., 2022). One way to address this limitation is to use tasks that assign monetary reward value to initially neutral stimuli, effectively rendering them salient and capable of competing for attentional resources. Indeed, prior research has shown that such value-associated, non-drug stimuli can capture attention even among healthy adult participants (Anderson et al., 2011, Bourgeois et al., 2016, Failing and Theeuwes, 2014, Meyer et al., 2020, Meyer et al., 2024). Given the association between adolescent alcohol use and reduced cognitive functioning on tests of visuospatial function, psychomotor speed, and cognitive control, attentional bias may be more pronounced during the early stages of such tasks, when attentional demands are highest (Sakai et al., 1998, Squeglia and Gray, 2016). Further investigating how alcohol use history affects attentional bias for non-drug reward stimuli may help uncover broader cognitive dysfunctions – particularly an impaired ability to shift attention away from salient but task-irrelevant information.

Both heightened reward sensitivity and poor executive function during adolescence are associated with heavier alcohol use in adolescence (Peeters et al., 2017, van Hemel-Ruiter et al., 2015). In turn, early and heavy drinking may further impair executive control, thereby driving involuntary attention and poor decision-making in the presence of salient cues (Koob and Volkow, 2016). Indeed, our lab has found that adults with a history of binge drinking before age 21 exhibited greater attentional bias not only toward alcohol-related cues but also toward non-drug, reward-associated stimuli (Elton et al., 2021).

Together, these findings highlight the potential long-term impact of adolescent alcohol use on both attentional and behavioral control processes in adulthood. Yet, despite evidence from animal studies and preliminary data in humans, there remains a critical gap in human research directly linking adolescent alcohol exposure to adult behavioral flexibility. To our knowledge, no studies have explored the potential interaction between adolescent and adult alcohol exposure in predicting behavioral inflexibility despite clear evidence from animal studies that this interaction is likely to exist. To address this gap, we used self-report measures of alcohol use alongside two behavioral tasks assessing different facets of behavioral flexibility. The first, the HABIT task, measures the ability to override previously well-learned stimulus-response associations in favor of newly learned ones, thereby indexing habitual behavior (McKim et al., 2016). The second, the Reward-Driven Attentional Bias (RDAB) task, assesses the extent to which non-drug reward-conditioned stimuli capture attention, based on a paradigm modified from Anderson (2011) and Failing and Theeuwes (2014). Notably, both tasks use non-drug stimuli, allowing us to examine general behavioral flexibility independent of alcohol-specific cues.

By administering two tasks that investigate distinct cognitive processes that both contribute to inflexible behavior, we aimed to capture complementary processes relevant to alcohol use disorder: rigid, habitual responding that characterizes drug-taking behavior, and heightened attention to reward-related cues that characterizes drug-seeking behavior. We hypothesized that adults with a history of heavy adolescent alcohol use would exhibit greater impairment in behavioral flexibility, reflected by greater habitual responding and attentional bias. We further predicted that these deficits would be exacerbated by heavier current alcohol use. Findings from this study aim to deepen our understanding of the origins of behavioral inflexibility as a potential intermediate phenotype of addiction.

## Materials and Methods

### General Procedure

Data were collected during two visits separated by at least one night of sleep. During the first visit, participants were trained in both the HABIT and RDAB tasks. During the second visit, participants were refreshed on both tasks prior to completing the testing portion of the HABIT task outside of the fMRI scanner (on a laptop) and the testing portion of the RDAB task while in the scanner.

### Participants

Healthy, right-handed participants (*n*=71 total) aged between 22 and 40 years (*M*= 26.7 ± 5.1) were recruited from the community via advertisements through email, social media, and flyers distributed around the University of North Carolina at Chapel Hill campus and surrounding area. Thirty-three subjects were recruited from a different study conducted in the lab in which the HABIT (McKim et al., 2016) was performed three different times under either sham or 10Hz transcranial alternating current stimulation prior to being recruited to the current study. Time since the last stimulation session was used as a covariate in all analyses. Subjects were recruited into two groups based on self-reported history of adolescent (before age 18) binge drinking or no adolescent binge drinking, with binge episode defined as ≥4 drinks/2 hr for females and ≥5 drinks/2 hr for males. Subjects were further stratified into groups based on whether they self-reported ≥2 binge episodes within the last 12 months or no binge episodes. Exclusion criteria included current use of psychoactive medication, psychiatric diagnosis within the past year (assessed via the Mini International Neuropsychiatric Interview; Sheehan et al., 1998), any neurological or systemic illness, any lifetime substance use disorder diagnosis (excluding AUD), and any contraindication to MRI. Each subject provided written informed consent as approved by the UNC Office of Human Research Ethics. We report associations between behavioral task performance and measures of GABA/glutamate balance in this sample elsewhere (Vidrascu et al., under review).

### Behavioral Inventories

Participants completed several standard questionnaires both in person and online via REDCap (Harris et al., 2009), a secure online platform for surveys and database management. These questionnaires covered self-reported substance use measured by the Alcohol Use Disorder Identification Test (AUDIT; Saunders et al., 1993) the Alcohol-Related Blackouts Questionnaire (ARBQ; Elton et al., 2021); the Carolina Alcohol Use Patterns Questionnaire (CAUPQ; Elton et al., 2021), the Customary Drinking and Drug Use Record (CDDR; Brown et al., 1998) and the Drinking Motives Questionnaire-Revised (DMQ-R; Cooper, 1994). Personality measures were assessed using the Barratt Impulsiveness Scale (Bis-11; Patton et al., 1995), The State-Trait Anxiety Index (STAI; Spielberger, 1985), the Conners’ Adult ADHD Rating Scales (CAARS; Conners et al., 1999), and the Locus of Control (LOC; Rotter, 1966). Two questionnaires measured behavioral flexibility; namely, the Value-Driven Attention Questionnaire (VDAQ; Anderson, 2020) and the Creature of Habit Scale (COHS; Ersche et al., 2017).

### Behavioral flexibility paradigms

#### Hidden Association between Images Task (HABIT)

The HABIT is a well-validated S-R learning task implemented in E-Prime 2.0 (PST Inc., Pittsburgh, PA) and is described in detail elsewhere (Boettiger and D’Esposito, 2005, McKim et al., 2016). Briefly, abstract stimuli representing a single square composed of multicolored blocks were briefly (for 700 ms) presented on a color LCD screen. Subjects were required to learn by trial and error to associate the block pattern with a press on an arbitrarily assigned button on a four-button keypad. During training, subjects learned to distinguish two sets (each consisting of two patterns) of S-R rules to a criterion of >90%. These well-learned sets are subsequently referred to as familiar (FAM) sets. Greater than 70% accuracy combined across both sets was needed to have successfully completed training. After >1 night’s sleep, participants returned to complete the testing portion of the HABIT. In Part 1, subjects were refreshed on the two FAM sets learned in session 1, and then 2-3 novel sets (NOV) were introduced in addition to control sets that consisted of unrelated stimuli. Following 6 “runs” of 15 blocks each (3 per block type), subjects were informed that the correct responses for 2 sets (one FAM and one NOV set) had changed (i.e., response devaluation), therefore requiring the new correct S-R associations to be learned through trial and error over 6 runs. This response devaluation manipulation allows habitual responding to be quantified when attempting to overcome both well-learned (FAM) and freshly learned (NOV) S-R associations. As a proxy for habitual action-selection we used perseverative errors, quantified as the percentage of incorrect responses (out of total) in which participants selected the previously correct button as opposed to selecting a new button when response contingencies were changed post-devaluation.

#### Reward-Driven Attentional Bias Task (RDAB)

The RDAB task has been extensively detailed elsewhere (Meyer et al., 2024) and consists of a reward conditioning training phase followed by an attention cueing paradigm testing phase. Briefly, in the initial reward conditioning task (Anderson et al., 2011), participants were presented with a series of circles on a laptop screen and were instructed to identify the orientation (vertical/horizontal) of a line within a target circle (blue/yellow) among an array of circles. One color was associated with a probabilistic (80%) monetary reward for accurate responses, while the other target color offered no monetary reward. The training phase comprised two runs of 60 trials each. We quantified performance using an inverse efficiency score (IES; RT/accuracy), to account for speed-accuracy tradeoffs (Statsenko et al., 2020) and compared performance between rewarded and unrewarded trials.

Subsequently, the testing phase of the RDAB task involved a visual spatial recognition task designed to assess attentional control in the presence of a reward-conditioned distractor (Failing and Theeuwes, 2014). During this phase, participants were presented with two colored circles on either side of a fixation cross. Each circle contained either a target letter (‘S’ or ‘P’) or a neutral letter (‘E’ or ‘H’). Participants were tasked with identifying the target letter while ignoring the colored circles, with correct responses not linked to any monetary reward. The testing phase comprised three runs, each consisting of 120 trials. Trial types were distinguished by the location of the previously conditioned stimulus relative to the target stimulus and included those in which the conditioned stimulus was located on the same (i.e., target-congruent, or “valid”) or opposite side (i.e., target-incongruent, or “invalid”) as the target stimulus, and trials in which no conditioned stimulus was present (i.e., “neutral”). Attentional bias measures were derived by comparing inverse efficiency scores (IES; RT/accuracy) for all trial types: *total bias* (invalid>valid), *facilitated attention* (valid>neutral), and *disengagement cost* (invalid>neutral). For both parts of the task, reaction times below 100 ms were excluded from further analyses and responses that were too slow (>800 ms) were coded as incorrect, consistent with previous studies utilizing these paradigms (Anderson et al., 2011, Failing and Theeuwes, 2014, Meyer et al., 2024).

#### Data analyses

##### Alcohol Use Composite Scores

To reduce the influence of recall bias regarding adolescent alcohol use (Livingston et al., 2016, Sartor et al., 2011), we accounted for shared variability across multiple related measures of alcohol consumption and consequences. A categorical principal components analysis was used to determine non-linear relationships between measures of alcohol use behavior and alcohol-related memory consequences (partial and total blackouts) during adolescence (before age 18) and within the past year (see Linting and van der Kooij, 2012 for review). Analyses were restricted to two dimensions, using oblimin rotation (Kaiser normalization) to allow components to rotate. Continuous measures of the first component for each alcohol use score, in addition to a binary measure, were used in subsequent analyses as noted. This approach was done for all subjects who completed the HABIT task (HABIT *group, n*=71) and again for the subset of participants who also completed the RDAB task (RDAB *group*, *n*=44). Participants were primarily recruited based on the presence or absence of adolescent binge drinking. To create two groups from our continuous principal component analysis, we applied a median split to the first component. For the HABIT group, a median split of the under 18 PCA component 1 (U18PC1) resulted in 36 participants in the *light adolescent use* group and 35 in the *heavy use* group. These groups did not differ in level of current alcohol use (χ^2^(1, *N*=71) = 2.5, *p=*.11). A median split of past year PCA component 1 (PYPC1) resulted in 35 participants in the *light past year use* group (24 *light* vs. 11 *heavy adolescent drinkers*) and 36 in the *heavy past year use* group (18 *light* vs. 18 *heavy adolescent drinkers*). In the RDAB Group, a median split of the under 18 PCA component 1 (U18PC1) resulted in 22 participants in the *light adolescent use* group and 23 in the *heavy adolescent use* group. These groups did differ in level of current alcohol use (χ^2^(1, *N*=45) = 6.41, *p=*.01). A median split of the past year PCA component 1 (PYPC1) resulted in 22 participants in the *light past year use* group (15 *light* vs. 7 *heavy adolescent drinkers*) and 23 in the *heavy past year use* group (7 *light* vs. 16 *heavy adolescent drinkers*).

Our main indices for inflexible behavior were perseverative responding post-contingency change in the HABIT task. We also used total attentional bias and its components (facilitated attention and disengagement cost) for previously rewarded trials in the RDAB task. In addition to using IES as our performance measure for attentional bias, we conducted similar analyses with reaction time and accuracy independently (see **Supplemental material**). Our *a priori* hypothesis was that adults with heavier current and adolescent alcohol use would exhibit greater inflexible behavior than those with lighter use. We first tested this hypothesis using discrete measures of current and adolescent alcohol use in two-way MANCOVA models with adolescent (light, heavy) and past year (light, heavy) alcohol use as fixed factors. We next investigated whether associations between current alcohol use and inflexible behavior were monotonic by using continuous measures of current alcohol use in multiple linear regression models with the covariates entered in Step 1, while adolescent alcohol use, past year alcohol use, and their interaction term were entered in order in Step 2.

Two extreme outliers were removed from analysis of the RDAB task. One participant exhibited unusually high performance on unrewarded trials during reward conditioning, far exceeding 12 (with a score of 29), which was the upper cutoff for outlier detection as determined with the interquartile range method. One participant exhibited unusually high facilitated attention, far exceeding −0.112 (with a score of −0.822), which was the lower cutoff for outlier detection. These outliers were also confirmed with visualization of boxplots and Q-Q plots. Consequently, 44 subjects remained for further analysis of the RDAB task.

All statistical analyses were conducted using SPSS v.29.0.1 (SPSS, Chicago, IL) and included time since last tACS session as a covariate for analysis of the HABIT task, and time since last drink containing alcohol for analysis of both tasks. There was no main effect of sex; as such, this was not included in final analyses. Significance testing was based on a bootstrapping procedure with 1000 samples and 95% CI. Group comparisons for demographic and psychometric variables were done using unpaired two-tailed *t*-tests for continuous measures and χ^2^ tests for categorical measures.

## Results

Demographic characteristics with alcohol use and psychometric measures are listed separately for participants who completed either only the HABIT task (**Tables 1 and 2**) or both the HABIT and RDAB tasks (Supplemental Tables 1 and 2). Study groups were well-matched for all demographic variables, with no significant differences in age, sex, race/ethnicity, or education. Subjects, on average, fell within the “low-risk consumption” band (AUDIT score<7), though scores varied based on level of alcohol use during adolescence (heavy adolescent drinkers, *M_AUDIT_ =* 6.1 ± 4.7; light adolescent drinkers, *M_AUDIT_* = 2.7 ± 2.4). Adults with a history of heavy adolescent alcohol use reported an earlier onset of first drink, drunk, and binge episode experience and a higher past year binge score (Townshend and Duka, 2002). Adults with light adolescent alcohol use had no binge drinking experience before age 18. See Supplemental Material for additional alcohol use and psychometric measure details.

**Table 1.**
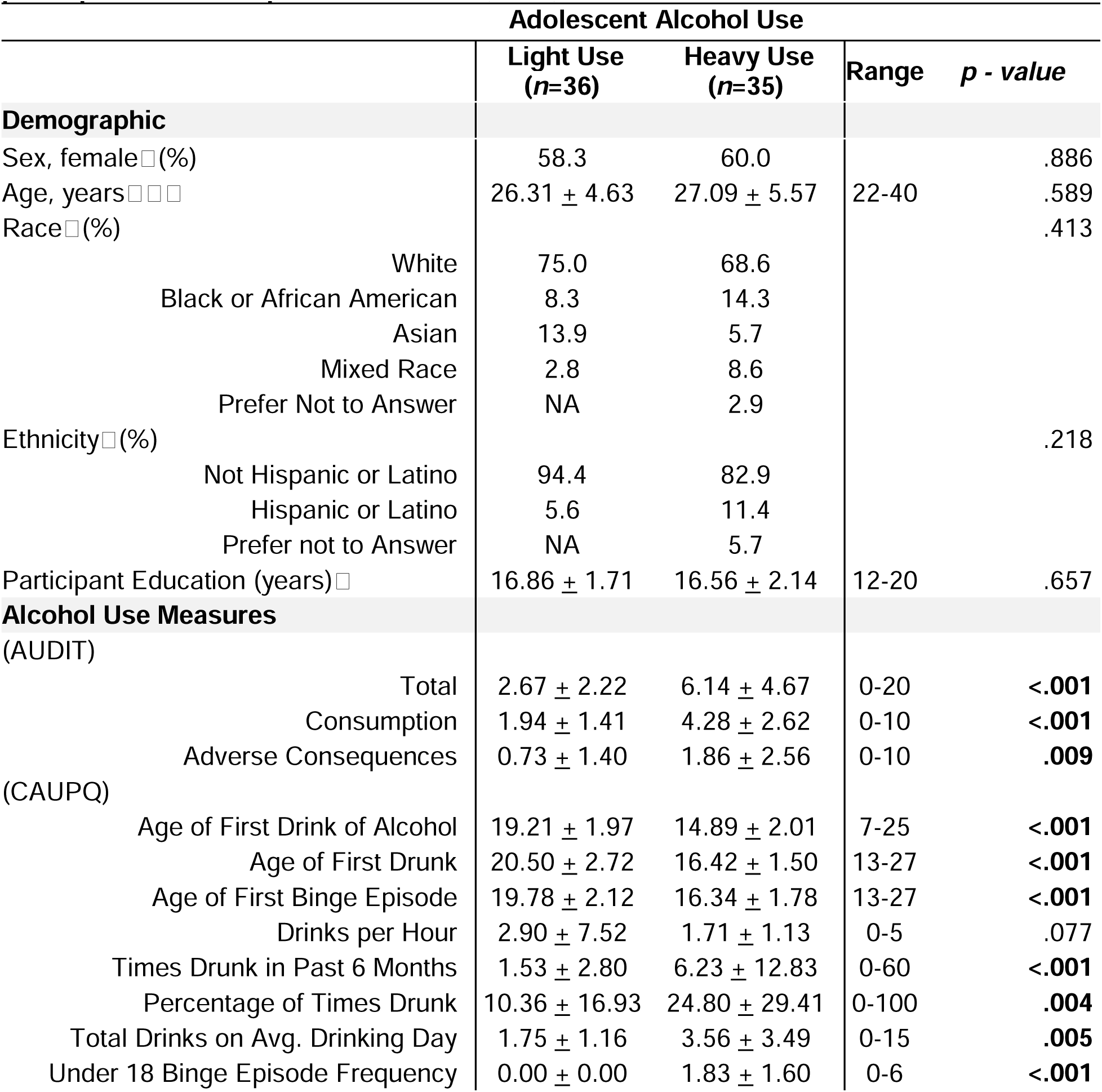
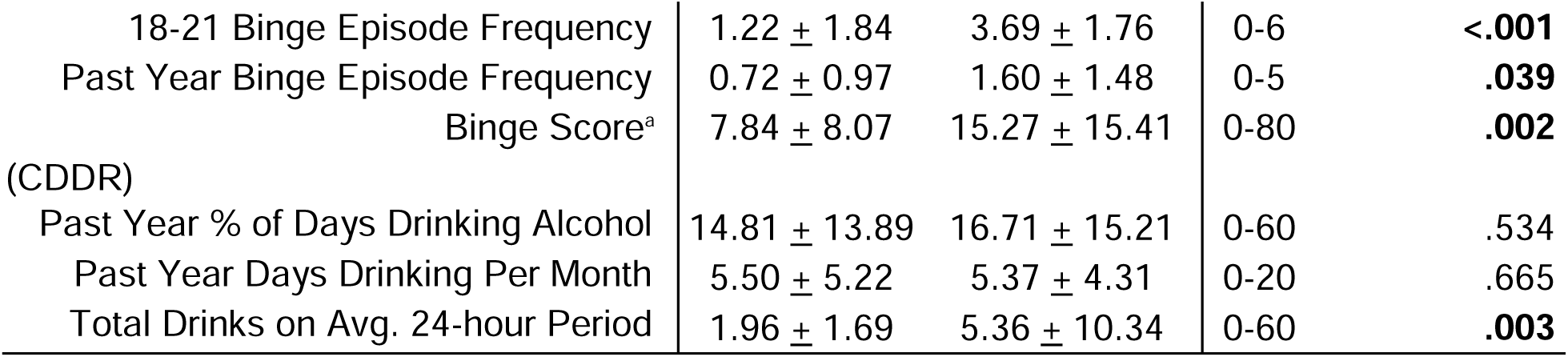
Demographic characteristics and descriptive statistics of alcohol use of participants who completed the HABIT (N=71)

**Table 2.**
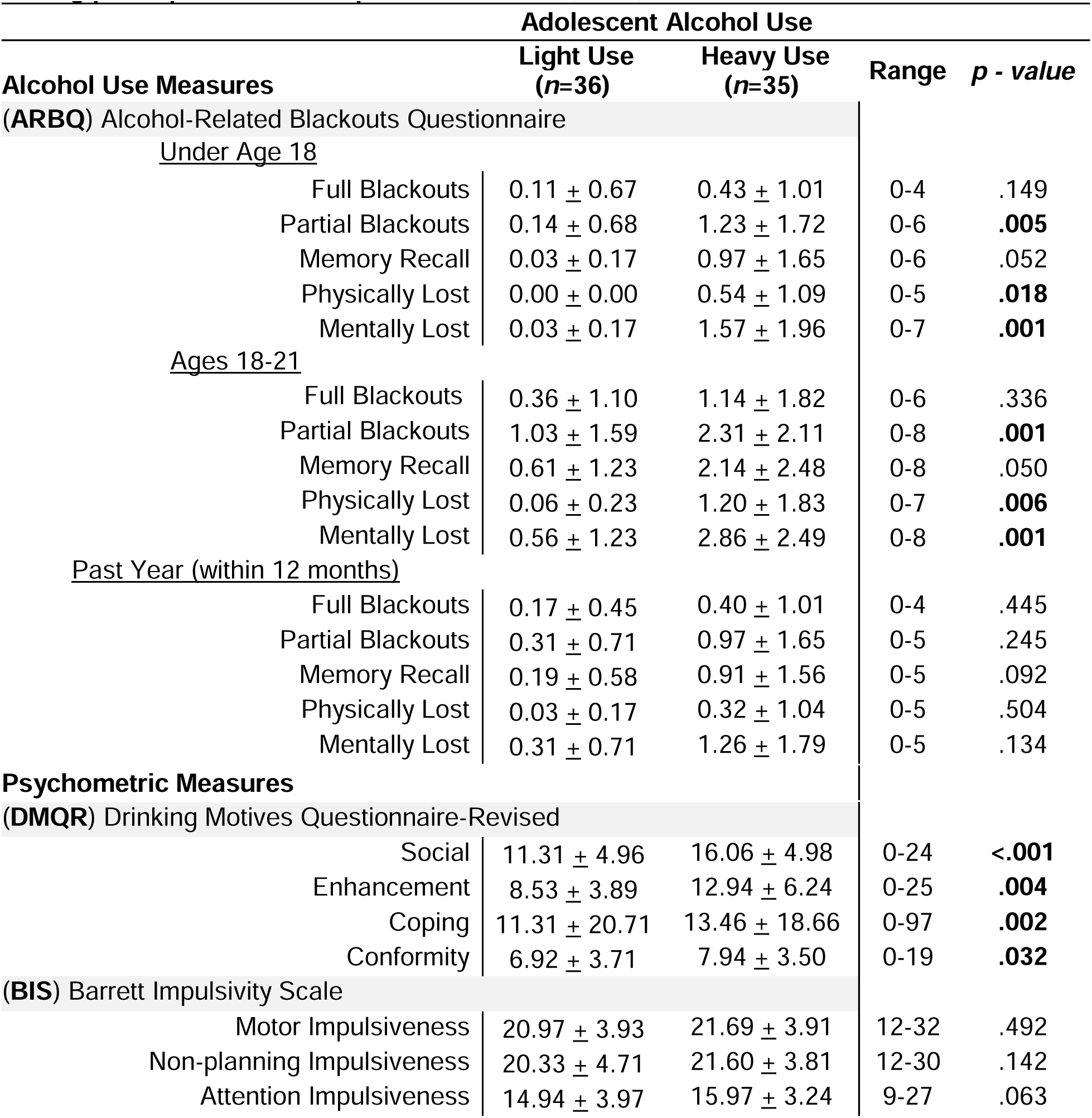
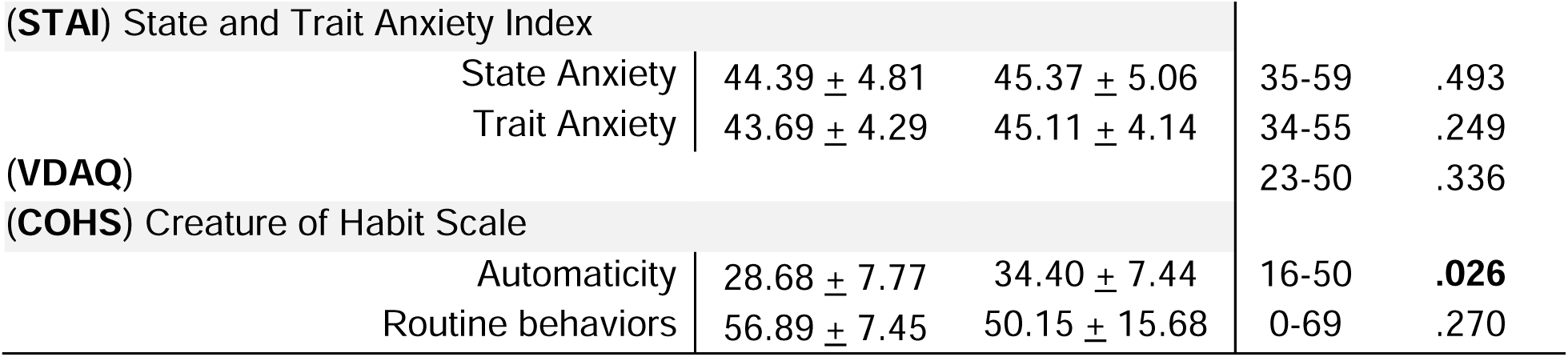
Descriptive statistics of alcohol/substance use and psychometric measures among participants who completed the HABIT (N=71)

### Alcohol use metrics

Composite scores were calculated for adolescent and past year alcohol use. Only variables with loadings of at least 0.5 were included in the final CAPTCA analysis. For group 1 (*n*=71), ninety-six percent of the variance across 9 variables describing alcohol use behavior and alcohol-related blackout before age 18 was captured in two components (Supplemental Table 3): U18PC1 explained 43% of the variability (Cronbach’s α = 0.892), and U18PC2 captured 40% of the variability (Cronbach’s α = 0.881). The variables which loaded most strongly onto U18PC1 described age of first drink, drunk, and binge drinking episode, and frequency of binge episodes before age 18. Since our *a priori* hypotheses were focused on behavioral differences as a function of binge drinking behavior, we used U18PC1 as our measure of adolescent alcohol use. Variables describing alcohol-induced blackout loaded most strongly onto U18PC2. We had 11 variables describing past year alcohol use and consequences, with sixty-nine percent of the variance captured in two components (Supplemental Table 4). The first component explained 35% of the variability (Cronbach’s α = 0.862), with significant loadings of five variables describing the quantity and frequency of alcohol consumption over the past 12 months. The second component explained an additional 34% of the variability (Cronbach’s α = 0.860) in the data, with six variables describing memory-induced blackout experiences, including instances of full and partial blackouts.

We found similar results for group 2 (*n*=44), but only 8 variables remained in the final CAPTCA analysis. Eighty-eight percent of the variance across 8 variables describing alcohol use behavior and alcohol-induced blackout before age 18 was captured in two components (Supplemental Table 5): U18PC1 explained 48% of the variability (Cronbach’s α = 0.873), and U18PC2 captured 33% of the variability (Cronbach’s α = 0.787). We found slightly different results for group 2 (*n=*44; Supplemental Table 6), such that only four variables loaded onto the component which described memory-induced blackout experiences (PYPC2), explaining 34% of the variability in the data (Cronbach’s α = .878). Total blackouts loaded onto the first component (PYPC1), with seven total variables explaining 42% of the variability (Cronbach’s α = .902).

### Alcohol Use and Habitual Action Selection

To evaluate the relationship between alcohol use and habitual action selection, we examined performance differences in the HABIT task; specifically, perseverative errors after outcome devaluation of a well-learned S-R association. First, we tested for group differences in the initial training session by running a repeated-measures ANCOVA model with the within-subjects factor being blocks to criterion for Pattern A and Pattern B. Adolescent (light, heavy) and past year (light, heavy) alcohol use were used as between-subject factors, and the covariates used were the time since last drink and the last tACS session. Regarding blocks to criterion, we did not find a significant effect of adolescent alcohol use (Wilks Lambda = 1.00, F_(1,64)_ =0.30, *p=*.58), but did find a significant effect of past year alcohol use (Wilks Lambda = 0.92, F_(1,66)_ =5.43, *p=*.02, □^2^ =.08), such that adults with heavy past year alcohol use learned slightly faster for Pattern B than adults with light use (M_heavy>light_= −0.50, *p=*.03, 95% CI -.949, -.054). Despite this slight difference in initial learning, there was no effect of adolescent or past year alcohol use on the execution of familiar sets or on the learning of novel sets during Test Part 1 (max *F*_(1,64)_=0.26, min *p=*.61). We then looked at group differences in perseverative errors made post devaluation phase during the HABIT Test Part 2, using a multivariate ANCOVA model with familiar and novel sets as dependent variables, and the same factors and covariates as described in prior models. We found a significant main effect of adolescent alcohol use on perseverative errors made for familiar sets (F_(1,65)_ =6.22, *p=*.02, □^2^ =.09), reflecting greater errors among those reporting heavy adolescent alcohol use (M_heavy>light_= 7.11, *p=*.015, 95% CI 1.415, 12.808; **Figure 1A**), but not for novel sets (F_(1,65)_ =0.94, *p=*.34, □^2^ =.01; **Figure 1B**). In contrast, we found no significant main effect of past year alcohol use on either set (max *F*_(1,65)_ =0.30, min *p=*.58), and no significant interaction effect of these two factors (max *F*_(1,65)_ =0.67, min *p=*.42).

**Figure 1.**
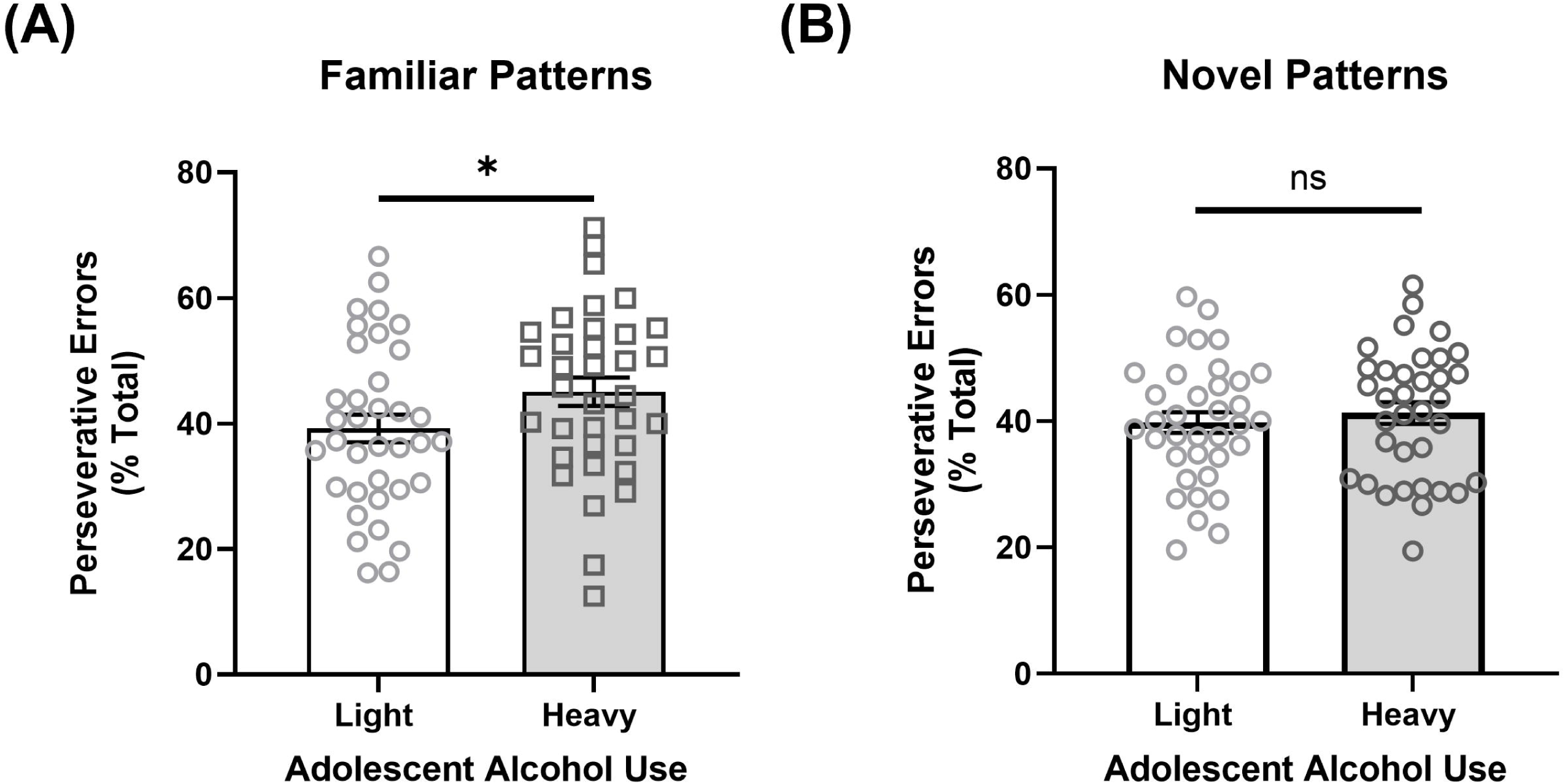
Perseverative responding after devaluation of well-learned and newly learned stimulus-response associations in the HABIT task between adults with light and heavy levels of adolescent alcohol use. A) Greater perseverative errors (out of total errors) made for well-learned patterns (familiar sets) among adults with a history of heavy adolescent alcohol use compared to adults with a history of light adolescent alcohol use (F_(1,65)_ =6.22, *p=*.015, □^2^ =.09); B) No difference in perseverative errors on newly learned patterns (novel sets) among adults with a history of either light or heavy adolescent alcohol use (F_(1,65)_ =0.94, *p=*.336, □^2^ =.01). \**p* < 0.05; Error bars represent standard error of the mean.

While we did not observe an interaction between adolescent and adult alcohol use in our MANCOVA analysis, we sought to examine if this interaction was observed when the potential monotonic relationship between current alcohol use and habitual behavior was preserved. We ran a linear regression model with covariates entered in Step 1, and adolescent alcohol use (binary), past year alcohol use (continuous), and their interaction term in Step 2. We found a significant predictive effect of heavy adolescent alcohol use (β*_standardized_*= .290, *p*=.01, 95% CI 1.78 18.51; **Table 3**). There was no significant effect of past year alcohol use (*p=*.29) and no significant interaction between these two variables (*p*=.70). Together, these results suggest that independent of current alcohol use behavior, adults with a history of heavy adolescent alcohol use exhibit greater perseverative behavior.

**Table 3.** Hierarchical Linear Multiple Regression Analysis of adolescent and past year alcohol use on perseverative errors post devaluation phase in the HABIT task (N=71)

|  |  |  |  |  |  |
| --- | --- | --- | --- | --- | --- |
| Variable | B | SE | $\beta$ | *p-value* | 95% CI |
| **Step 1- covariates** |  |  |  |  |  |
| Time since last drink | -.002 | .007 | -.015 | .717 | [-.013, .013] |
| Time since last tACS session | 12.780 | 2.908 | .475 | <.001 | [6.838, 17.917] |
| **Step 2 - predictors** |  |  |  |  |  |
| Heavy adolescent alcohol use | 7.787 | 2.943 | .290 | .008* | [1.782, 18.513] |
| Past year alcohol use | -2.598 | 2.672 | -.194 | .292 | [-7.305, 3.649] |
| Adolescent X Past year alcohol use | 1.156 | 3.282 | .075 | .701 | [-6.172, 6.948] |
| *Note:* Heavy adolescent alcohol is a binary variable; 0, light use; 1, heavy use.<br>B, unstandardized coefficient; $\beta$ , standardized coefficient; SE, standard error. *p-value* based on 1000 bootstrap samples. * *p* <0.05 | | | | | |

### Alcohol Use and Reward Conditioning

We first assessed the discrepancy in performance (IES) for rewarded and unrewarded trials for the entire sample in the training phase of the RDAB task by using a paired samples *t-*test and found that participants performed significantly better on trials with the rewarded versus unrewarded circle (*M*_R>U_= −0.573, t=-4.48, p<0.001, Cohen’s d=-.683; **Figure 2**). While this is not a measure of behavioral inflexibility, it does form the basis for the subsequent task and indexes reward-driven attentional capture in a goal-congruent setting. As such, we explored the impact of alcohol use on performance. No significant main effects of adolescent alcohol use were found when looking at differences in RT and accuracy separately (Supplemental Table 7). We next utilized a two-way ANCOVA with adolescent (light, heavy) and past year (light, heavy) alcohol use as fixed factors and the difference between rewarded versus unrewarded (R>U) IES as the dependent variable, while controlling for time since last drink containing alcohol. The main effect of adolescent and past year alcohol use, as well as the interaction of these two factors on reward conditioning (max *F*_(1,38)_ =0.74, min *p=*.39), were non-significant. When examining monotonic associations between past year alcohol use and performance in regression analyses, we similarly found no evidence for an effect of past year or adolescent exposure, or an interaction between these variables, in predicting reward-driven attentional capture in a goal-congruent task (min *p*=.25; Supplemental Table 8). These findings suggest that neither adolescent nor recent alcohol use significantly influences reward-driven attentional capture in a goal-congruent setting.

**Figure 2.**
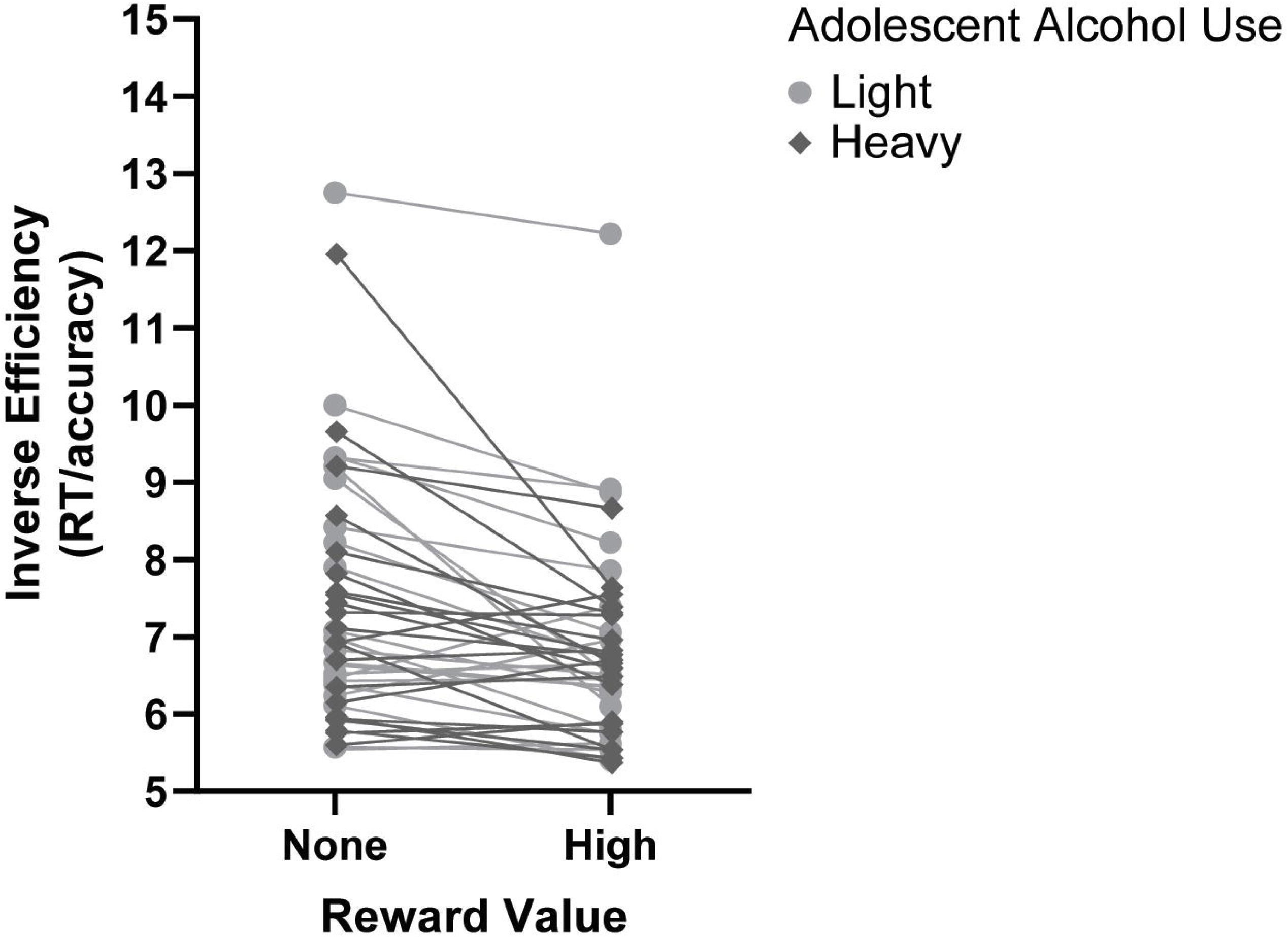
Performance for rewarded and unrewarded trials during the RDAB task reward conditioning phase for the entire sample (N=44) Participants performed significantly better on trials with the rewarded versus unrewarded circle, as reflected by a lower inverse efficiency score (*M*_R>U_= −0.573, t=-4.48, p<0.001, Cohen’s d=-.683). Inverse efficiency (IE) = reaction time/accuracy. Variability in performance is shown between adults with a history of light and heavy adolescent alcohol use, although no significant main effect was found.

### Alcohol Use and Attentional Bias

Adolescent group differences in RT, accuracy, and IES across all three runs for all trial types are shown in Supplemental Tables 9 and 10. Independent of alcohol use, we first looked at differences in attentional bias, our other metric of behavioral inflexibility, during the early phase (i.e., run 1) wherein attentional demands may be greatest and found a significant difference in all three measures using the Friedman test (□^2^=17.17, *p*<.001, df=2; **Figure 3A**). A Wilcoxon signed-rank test showed that total bias did not significantly differ from facilitated attention (*Z*=-1.30, *p=*0.19), but disengagement cost differed from both total bias (*Z=*-3.76, *p<*0.001) and facilitated attention (*Z=*-4.10, *p<*0.001). To evaluate the relationship between alcohol use and attentional bias, we used a two-way MANCOVA with adolescent (light, heavy) and past year (light, heavy) alcohol use as fixed factors, while controlling for time since last drink containing alcohol. There was no significant main effect of adolescent alcohol use on any of the three performance measures (max *F*_(1,36)_=1.39, min *p=*.25). We did not find a significant main effect of past year alcohol use on total bias or facilitated attention (max *F*_(1,36)_=0.96, min *p=*.33), but found a marginally significant effect on disengagement cost (F_(1,36)_=3.57, *p=*.07, □^2^ =.090). We found no significant interaction effect of adolescent and past year alcohol use any performance measure (max *F*_(1,36)_=0.72, min *p=*.40).

**Figure 3.**
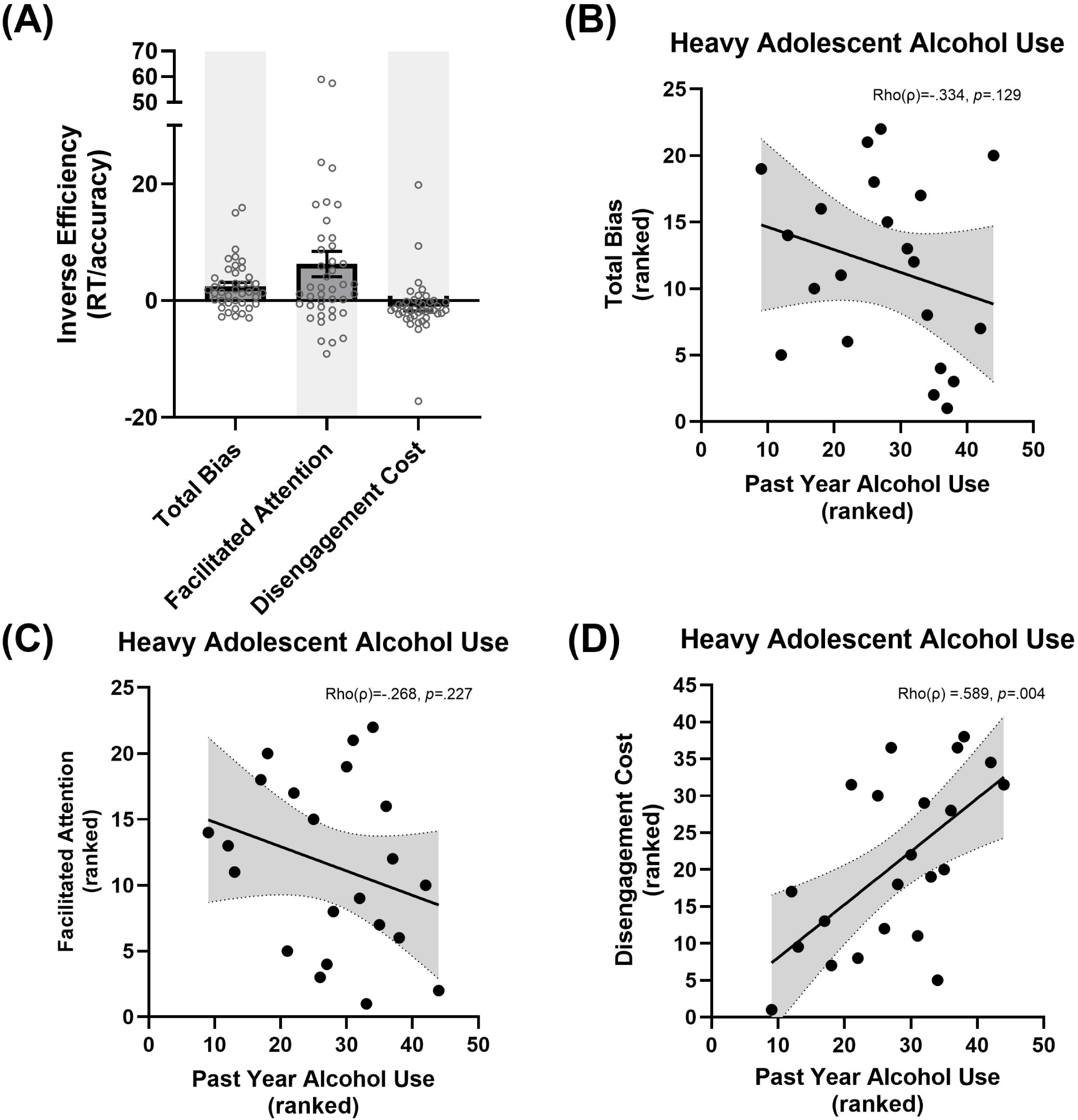
Relationship between adolescent and current alcohol use and attentional bias in the RDAB task. A) A Wilcoxon signed-rank test showed that total bias did not significantly differ from facilitated attention (*Z*=-1.30, *p=*0.19), but disengagement cost differed from both total bias (*Z=*-3.76, *p<*0.001) and facilitated attention (*Z=*-4.10, *p<*0.001). B) Among adults with a history of heavy adolescent alcohol use, we found no significant association between past year alcohol use and total bias (Rho’s ρ=-0.334, *p*=.13) or C) facilitated attention (Rho’s ρ=-0.268, *p*=.23), but did find a significant association with D) disengagement cost (Rho’s ρ=0.589, *p*=.004). Analyses controlled for recency of alcohol consumption. 95% confidence interval bands shown.

As above, we next ran separate linear regression models to examine monotonic associations between past year alcohol use and behavioral inflexibility. We included time since last drink in Step 1, and individual measures of adolescent (binary) and past year alcohol use (continuous), in addition to their interaction term, in Step 2. We did not find any significant predictive effects of adolescent or past year alcohol use, nor their interaction, on facilitated attention (min *p=*.65) or total bias (min *p=*.23). We found a significant predictive effect of adolescent alcohol use on disengagement cost (β*_standardized_*= -.526, *p*=.042, 95% CI −26.449 −2.942), as well as an interaction effect of adolescent and past year alcohol use (β*_standardized_*= .601, *p*=.040, 95% CI .094 .938; **Table 4**). To decompose this significant interaction, we used a Spearman correlation. Among adults with heavy adolescent alcohol use, we did not find a significant correlation between past year alcohol use and either total bias (Rho’s ρ=-0.334, *p*=.13; **Figure 3B**) or facilitated attention (Rho’s ρ=-0.268, *p*=.23; **Figure 3B**), but found a significantly positive association with disengagement cost (Rho’s ρ=0.589, *p*=.004; **Figure 3D**). Thus, greater past year alcohol use was associated with increased difficulty disengaging from a reward-related distractor, specifcially among individuals with a history of heavy adolescent alcohol use.

**Table 4.**
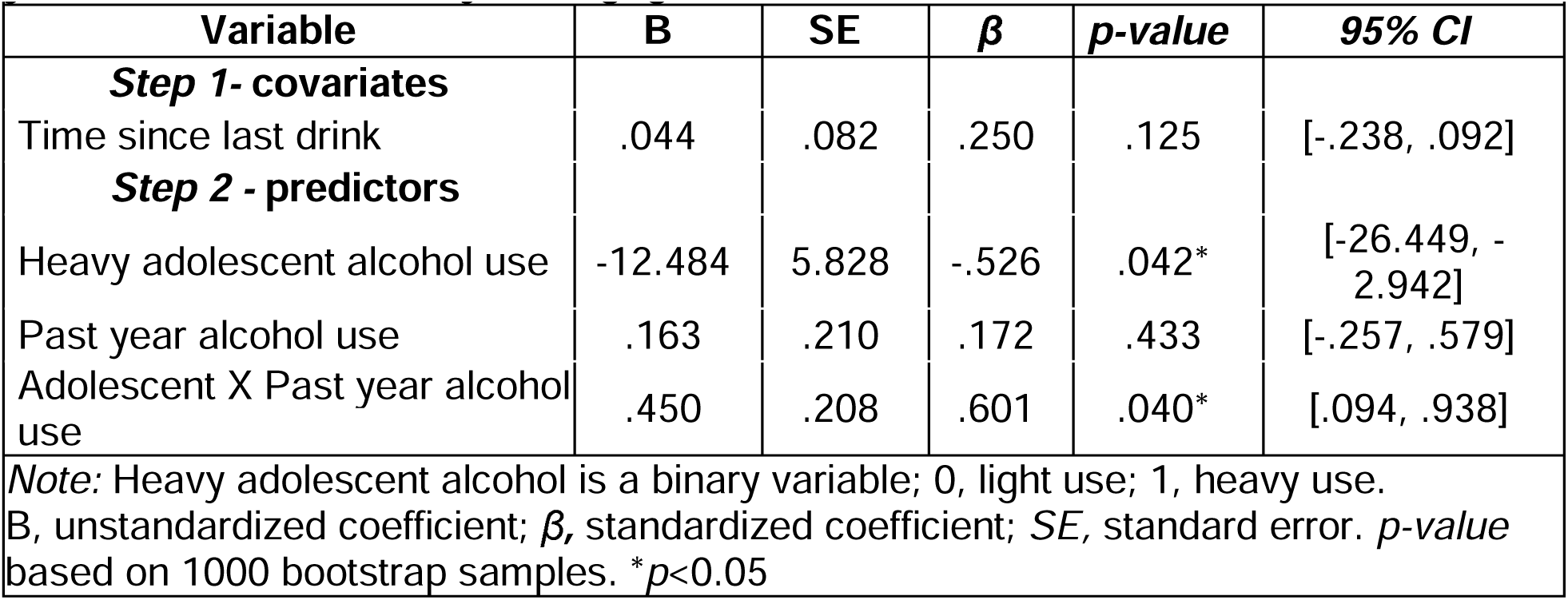
Hierarchical Linear Multiple Regression Analysis of adolescent and past year alcohol use on early disengagement cost in the RDAB task (N=44)

## Discussion

To the best of our knowledge, this is the first human study to present preliminary evidence linking adolescent alcohol use to reduced behavioral flexibility in adulthood. Specifically, we found that adults who reported binge drinking during adolescence were more likely to rely on habitual rather than goal-directed action, regardless of their current drinking behavior. Furthermore, among individuals with a history of adolescent drinking, greater alcohol use in the past year was associated with an increase in disengagement cost, reflecting difficulty in overcoming a reward-related distractor. These findings suggest that different aspects of behavioral flexibility, such as habitual responding and attentional disengagement, may be influenced in distinct ways by early alcohol exposure and, in some cases, continued use. Longitudinal studies are needed to determine whether these cognitive patterns are lasting consequences of adolescent drinking or risk factors that predate both adolescent and current use.

Using the HABIT paradigm, we found that adults with a history of adolescent binge drinking exhibited greater difficulty overriding habitual responses, independent of their current alcohol use. This study provides robust evidence that alcohol use before age 18 may disrupt the balance between goal-directed and habitual control systems. Although our measure of adolescent alcohol use relied on retrospective self-report and is therefore subject to recall bias (Livingston et al., 2016, Sartor et al., 2011), the composite score we used emphasized early-onset markers of alcohol exposure – such as age of first drink, first drunkenness, and first binge episode – as well as the frequency of binge drinking. These results are consistent with rodent models showing the persistent effects of adolescent alcohol exposure on behavioral flexibility, including set-shifting ability and reversal learning (Dannenhoffer et al., 2021, Gomez et al., 2021). While further longitudinal evidence is necessary, these findings suggest that adolescent alcohol exposure may increase risk for future psychopathology, including future AUD, by increasing a tendency towards habitual responding.

Unlike other set-shifting tasks, a methodological strength of the HABIT task is that it directly assesses the ability to override well-learned stimulus-response (S-R) associations in favor of newly learned ones (Boettiger and D’Esposito, 2005, McKim et al., 2016, Uddin, 2021). This feature makes it particularly well-suited for studies examining behavioral inflexibility as a potential intermediate phenotype of addiction, which is characterized by progression from goal-directed to habitual alcohol use (for review see Barker and Taylor, 2014). An additional advantage of the HABIT task is that it minimizes working memory demands during the test phase by requiring participants to overlearn initial S-R contingencies during training. Furthermore, by incorporating both familiar and novel S-R pairings in the test phase, the task enables a more precise dissociation of habitual responding from other executive functions – such as working memory, response inhibition, and attentional control. While each of these executive functions has been implicated in addiction (Koob and Volkow, 2016), isolating habitual responding as a distinct cognitive mechanism provides a more targeted behavioral marker.

While we observed interactions between adolescent and adult alcohol use in predicting reward-driven attentional capture when reward was not goal-congruent, we did not observe a similar effect on attentional engagement with these cues when reward was goal-congruent. While enhanced engagement with a conditioned cue may reflect goal-directed, facilitated attention, difficulty disengaging suggests impaired attentional control and a potential shift towards more automatic, or inflexible, habitual responding. Supporting this interpretation, we recently showed (Meyer et al., 2024) that heightened activity in the striatum during reward training was associated with greater subsequent recruitment of the anterior cingulate cortex in the presence of the reward-related distractor, a region important for exerting self-control and monitoring conflict (Tang et al., 2015). This suggests that shifting attention away from rewarding cues requires increased cognitive effort and control, with chronic alcohol use increasing such demands. Future studies can further investigate how this neural circuitry is shaped by patterns of alcohol use across adolescence and adulthood.

Interestingly, we found that current alcohol use exacerbated the effects of adolescent alcohol use on attentional bias, but not on habitual action-selection. Adolescent alcohol use alone was not clearly associated with attentional bias, suggesting that more recent and repeated alcohol exposure may be necessary to elicit measurable alterations in attentional salience (Bollen et al., 2022). While our analysis controlled for the recency of alcohol consumption, individuals with heavier drinking patterns – characterized by both frequency and quantity – may undergo more pronounced changes in the neural circuitry responsible for detecting and responding to salient stimuli. Consistent with this interpretation, previous work from our lab (Elton et al., 2021) found that adult social drinkers with a history of adolescent alcohol use before age 21 showed increased attentional bias toward both alcohol- and reward-related cues. Moreover, although current alcohol use was not directly examined, the observed bias was mediated by functional connectivity between brain regions involved in processing and communicating reward salience (Ghahremani et al., 2015, Wang et al., 2015, Uddin, 2021). Our current findings, however, suggest that drinking behavior before age 18 may be sufficient to produce long-lasting changes in attentional bias. More specifically, we demonstrated that while alcohol use history is not associated with differences in learning reward-related attentional bias, it is associated with sustained difficulty in disengaging from reward-related distractors even when at odds with current goals and no longer signalling reward. Other research has demonstrated that heightened sensitivity to reward among adolescents, particularly those around age 15, contributes to greater attentional bias toward alcohol-related cues (van Hemel-Ruiter et al., 2015). Collectively, these findings suggest that continued drinking into adulthood may further sensitize dopamine-related neural circuits, amplifying attentional responses to cues associated with reward motivation (Berridge and Robinson, 2016). However, because adults with heavy adolescent alcohol use initiated drinking earlier in adolescence, the current study cannot clearly disentangle specific developmental effects of alcohol from overall duration of use. To better understand the mechanisms of these effects, future studies can utilize longitudinal designs to examine how early alcohol use influences attentional bias in adulthood – particularly toward non-drug related cues – and to delineate the neurobiological processes underlying its attentional components.

A limitation of the present analysis is that the relatively small sample size likely reduced our ability to detect more nuanced effects and interactions. For instance, although we observed no main effect of sex on task performance, it is possible that an interaction between sex and alcohol use would emerge with a larger sample, as prior animal research has demonstrated sex-specific effects of alcohol exposure on reward conditioning and flexibility tasks, such as set-shifting (Madayag et al., 2017, Varlinskaya et al., 2020). Another limitation relates to our measure of adolescent alcohol use, which captured drinking behavior prior to age 18. While our data show that adults with a history of heavy adolescent alcohol use also had greater binge drinking frequency between ages 18-21, we did not investigate individual differences during this later developmental window. Animal studies suggest that the timing of alcohol exposure within adolescence can differentially affect various outcomes (Spear, 2015, Varlinskaya et al., 2020). Given the high prevalence of binge drinking among young adults and its documented neurocognitive consequences (Carbia et al., 2018, Hermens and Lagopoulos, 2018, Perez-Garcia et al., 2022), it is possible that we missed to account for additional variability in behavioral inflexibility. Future studies can examine how alcohol exposure during different stages of adolescence and early adulthood may impact behavioral flexibility.

In summary, our findings suggest that adolescent and current alcohol use may contribute differently to behavioral inflexibility in adulthood. Specifically, adolescent but not adult use was associated with habitual responding, whereas adult and adolescent use both contributed to inflexible attention towards reward. This study provides a foundation for future research exploring the shared and distinct mechanisms underlying a potential intermediate phenotype of addiction: behavioral inflexibility. Longitudinal studies will be essential to clarify how binge drinking at different development stages – early in life versus later in adulthood – affects neural circuitry and contributes to the risk of developing an alcohol use disorder.

## Data Availability

All data produced in the present study are available upon reasonable request to the authors

## Acknowledgements

This work was supported by Award Numbers P60AA011605 (CAB & DLR), T32DA007244 (EMV & MMR), T32AA007573 (EMV), and F31AA028427 (MMR) from the National Institutes of Health.

