## Supplemental Material for "Linking Adolescent Alcohol Use to Adult Behavioral Flexibility: Habitual Action-Selection and Attentional Bias"

**1	Reward-Driven Attentional Bias (RDAB) Task Behavioral Results for Reaction Time and Accuracy Separately**

Previous studies have reported reaction time and accuracy as their measures of attentional bias. Although we chose to use the inverse efficiency score as our measure to account for speed and accuracy tradeoff, below we provide results from the reward conditioning and cueing task separately for reaction time and accuracy as a function of adolescent alcohol use (Supplemental Tables 7, 9, and 10).

1.1 **Reward Conditioning**

To assess the effects of reward on training, we used a 2x2 repeated-measures ANOVA with reward status (rewarded, unrewarded) and session as within-subjects factors and adolescent alcohol use (light, heavy) as a between-subjects factor, controlling for past year alcohol use. This was done separately for RT and accuracy (Supplemental Table 7). With regards to RT, there was a main effect of reward type (*F*_(1, 41)_=21.65, *p*<.001, *η*^2^=.35), such that participants were faster to the rewarded than the unrewarded color. There was also a main effect of session (*F*_(1, 41)_=9.84, *p*=.003, *η*^2^=.19), wherein participants’ RT were faster in Session 2 than in Session 1. There was also a reward by session interaction (*F*_(1, 41)_=29.28, *p*<.001, *η*^2^=.42), such that in Session 2, participants were faster to the rewarded than the unrewarded color. We also found an interaction effect between session and adolescent alcohol use (*F*_(1, 41)_=11.58, *p*=.001, *η*^2^=.22), wherein adults with a history of heavy adolescent alcohol were faster during Session 1 than adults with a history of light adolescent alcohol use. For accuracy, there was a main effect of reward (*F*_(1, 41)_=18.81, *p*<.001, *η*^2^=.31), such that participants were more accurate for the rewarded relative to unrewarded color. There was no main effect of session (*p=*.09), but there was an interaction effect between reward and session (*F*_(1, 41)_=5.15, *p*=.03, *η*^2^=.11), wherein participants were more accurate for the rewarded color during Session 2 compared to Session 1.

1.2 **Attention Cueing Paradigm**

We used repeated measures ANOVA with cue type (invalid, neutral, valid) as the within-subjects factor and adolescent alcohol use (light, heavy) as the between-subjects factor. This was done separately for RT and accuracy (Supplemental Table 9). With regards to RT, there was a main effect of cue type (*F*_(1, 41)_=12.68, *p*<.001, *η*^2^=.24). Within-subjects contrasts revealed a quadratic effect, such that participants were faster for neutral cues than for both invalid and valid cues. There was no interaction effect between cue type and adolescent alcohol use (*p*=.26). For accuracy, within-subjects contrasts also revealed a quadratic effect of cue type (*F*_(1, 41)_=12.86, *p*<.001, *η*^2^=.24), such that participants were more accurate for neutral cues than for invalid or valid cues. There was no interaction effect between cue type and adolescent alcohol use (*p=.*95).

**2	Behavioral Results for Unrewarded Trials during Attention Cueing Paradigm**

Follow-up analyses were conducted for previously unrewarded trials to determine whether group differences in attentional bias were specific to reward history or general learning experience. Adolescent group differences in RT, accuracy, and IES are shown in Supplemental Table 10. We first looked at whether there were any group differences in all 3 performance measures using a two-way MANCOVA with adolescent (light, heavy) and past year (light, heavy) alcohol use as fixed factors, while controlling for time since last drink containing alcohol. We did not find main effects of adolescent (Wilks Lambda = 0.96, F_(1,37)_=0.54, *p=*.66) or past year alcohol use (Wilks Lambda = 0.90, F_(1,37)_=1.42, *p=*.25), nor any interaction effect of these two factors (Wilks Lambda = 0.97, F_(1,37)_=0.44, *p=*.73) across attentional bias performance measures. There was no main effect of adolescent alcohol use on any of the three performance measures (max *F*_(1,39)_=0.47, min *p=*.50), nor a main effect of past year alcohol use (max *F*_(1,39)_=2.59, min *p=*.12), or any interaction effect (max *F*_(1,39)_=0.44, min *p=*.51).

Next, we looked at performance for each attentional bias measure across all three runs of the task using a repeated-measures two-way MANCOVA with run (early, mid, late) as the within-subjects factor. We found no violation of the Mauchly’s Test for Sphericity for any of the attentional bias measures (min *p=.*720). We first looked at total bias across the three runs of the task and did not find within-subject effects of adolescent alcohol use (Wilks Lambda = 0.91, F_(2,35)_=1.64, *p=*.21), but did find effects of past year alcohol use (Wilks Lambda = 0.74, F_(2,35)_=6.01, *p=*.01, ɳ^2^*_p_*=.256). Specifically, we found a significant quadratic-orienting effect in performance (F_(1,36)_=9.80, *p=*.003, ɳ^2^*_p_*=.214), such that adults with heavy past year alcohol use exhibited greater total bias during run 2 than during run 1 (M_run2-run1_=9.16, *p=*.006), whereas adults with light past year alcohol use exhibited less total bias during run 2 than during 1 (M_run2-run1_=-7.51, *p=*.036). Adults with heavy past year alcohol use also exhibited greater total bias during run 2 than adults with light use (M_heavy-light_=13.80, *p=*.001). We then looked at disengagement cost and did not find within-subject effects of adolescent (Wilks Lambda = 0.92, F_(2,35)_=1.63, *p=*.21) or past year alcohol use (Wilks Lambda = 0.89, F_(2,35)_=2.15, *p=*.13), or any interaction effect (Wilks Lambda = 0.98, F_(2,35)_=0.38, *p=*.68). We did not find a main effect of adolescent or past year alcohol use, or any interaction effect (max *F*_(2,36)_=2.28, min *p=*.14). Last, we looked at facilitated attention across the three runs of the task and did not find within-subject effects of adolescent (Wilks Lambda = 1.00, F_(2,35)_=0.43, *p=*.96) or past year alcohol use (Wilks Lambda = 0.99, F_(2,34)_=0.23, *p=*.80), or any interaction effect (Wilks Lambda = 0.91, F_(2,35)_=1.69, *p=*.20). We did not find a main effect of adolescent or past year alcohol use, or any interaction effect (max *F*_(2,36)_=1.96, min *p=*.15).

We further probed the interacting effects of adolescent and past year alcohol use on attentional bias measures for outcome variables that we found were significant for previously rewarded trials. No significant Spearman bivariate correlations were found between past year alcohol use and outcome variables among adults with light (min *p=*.080) or heavy (min *p=*.093) adolescent alcohol use. Further, regression models indicated no significant main or interaction effects of adolescent and past year alcohol use on early disengagement cost (min *p=.*298).

**3	Supplementary Tables**

| **Supplemental Table 1. Demographic characteristics and descriptive statistics of alcohol use of participants who completed the HABIT and RDAB (N=44).** | | | | | |
| --- | --- | --- | --- | --- | --- |
|  | **Adolescent Alcohol Use** | |  |  | |
|  | **Light Use (*n*=22)** | **Heavy Use (*n*=23)** | **Range** | | ***p - value*** |
| **Demographic** |  |  |  | |  |
| Sex, female (%) | 63.6 | 65.2 |  | | .912 |
| Age, years | 25.95 ± 4.99 | 26.39 ± 5.48 | 22-40 | | .781 |
| Race (%) |  |  |  | | .367 |
| White | 68.2 | 60.9 |  | |  |
| Black or African American | 4.5 | 13.0 |  | |  |
| Asian | 22.7 | 8.7 |  | |  |
| Mixed Race | 4.5 | 13.0 |  | |  |
| Prefer Not to Answer | NA | 4.3 |  | |  |
| Ethnicity (%) |  |  |  | | .068 |
| Not Hispanic or Latino | 100 | 78.3 |  | |  |
| Hispanic or Latino | NA | 13.0 |  | |  |
| Prefer not to Answer | NA | 8.7 |  | |  |
| Participant Education (years) | 16.86 ± 1.78 | 16.68 ± 1.91 | 12-20 | | .486 |
| **Alcohol Use Measures** |  |  |  | |  |
| (AUDIT) |  |  |  | |  |
| Total | 2.45 ± 2.24 | 4.82 ± 4.22 | 0-20 | | **.014** |
| Consumption | 1.64 ± 1.22 | 3.50 ± 2.22 | 0-10 | | **.001** |
| Adverse Consequences | 0.82 ± 1.59 | 1.32 ± 2.40 | 0-10 | | .211 |
| (CAUPQ) |  |  |  | |  |
| Age of First Drink of Alcohol | 19.80 ± 2.04 | 15.14 ± 2.53 | 7-25 | | **<.001** |
| Age of First Drunk | 20.80 ± 2.27 | 17.05 ± 1.84 | 13-27 | | **<.001** |
| Age of First Binge Episode | 21.92 ± 2.91 | 16.84 ± 1.43 | 13-27 | | **<.001** |
| Drinks per Hour | 1.03 ± 0.80 | 1.65 ± 1.13 | 0-5 | | **.049** |
| Times Drunk in Past 6 Months | 1.32 ± 2.95 | 5.14 ± 12.46 | 0-60 | | **.001** |
| Percentage of Times Drunk | 11.55 ± 20.33 | 20.91 ± 24.99 | 0-100 | | **.028** |
| Total Drinks on Avg. Drinking Day | 1.50 ± 0.96 | 3.00 ± 2.30 | 0-15 | | **.001** |
| Under 18 Binge Episode Frequency | 0.00 ± 0.00 | 1.59 ± 1.74 | 0-6 | | **.004** |
| 18-21 Binge Episode Frequency | 0.82 ± 1.74 | 3.17 ± 2.08 | 0-6 | | **.001** |
| Past Year Binge Episode Frequency | 0.50 ± 0.86 | 1.36 ± 1.33 | 0-5 | | .132 |
| Binge Score^a^ | 7.75 ± 9.26 | 11.36 ± 6.85 | 0-80 | | **.022** |
| (CDDR) |  |  |  | |  |
| Past Year % of Days Drinking Alcohol | 11.28 ± 13.73 | 15.35 ± 11.86 | 0-60 | | .084 |
| Past Year Days Drinking Per Month | 4.05 ± 4.45 | 5.91 ± 4.82 | 0-20 | | .056 |
| Total Drinks on Avg. 24-hour Period | 1.64 ± 1.05 | 6.04 ± 12.44 | 0-60 | | **.004** |

Values are reported as mean ± SD. Reported *p-*values reflect the results of unpaired two-tailed comparison between groups: Mann-Whitney U non-parametric test for continuous measures or χ^2^ tests for categorical measures. Boldface indicates a significant difference between groups at *p*<0.05. AUDIT, Alcohol Use Disorders Identification Test; CAUPQ, Carolina Alcohol Use Patterns Questionnaire. CDDR,  Customary Drinking and Drug Use Record.

*Townshend and Duka 2002 binge score.

(CAUPQ) *Binge Episode Frequency*: 1, Never; 2, 1-3x; 3, 4-6x; 4, 7-12x; 5, 2-3x/month; 6, Weekly; 7, >1x/week

| **Supplemental Table 2. Descriptive statistics of alcohol/substance use and psychometric measures among participants who completed the HABIT and RDAB (N=44)** | | | | |
| --- | --- | --- | --- | --- |
|  | **Adolescent Alcohol Use** | |  |  |
| **Alcohol Use Measures** | **Light Use**  **(*n*=22)** | **Heavy Use**  **(*n*=23)** | **Range** | ***p - value*** |
| (**ARBQ**) Alcohol-Related Blackouts Questionnaire | | |  |  |
| Under Age 18 |  |  |  |  |
| Full Blackouts | 0.18 ± 0.85 | 0.13 ± 0.63 | 0-4 | .368 |
| Partial Blackouts | 0.23 ± 0.87 | 0.74 ± 1.32 | 0-6 | .394 |
| Memory Recall | 0.00 ± 0.00 | 0.96 ± 1.46 | 0-6 | .085 |
| Physically Lost | 0.00 ± 0.00 | 0.26 ± 0.62 | 0-5 | .122 |
| Mentally Lost | 0.00 ± 0.00 | 0.96 ± 1.46 | 0-7 | **.029** |
| Ages 18-21 |  |  |  |  |
| Full Blackouts | 0.41 ± 1.33 | 0.78 ± 1.54 | 0-6 | .469 |
| Partial Blackouts | 1.09 ± 1.88 | 2.00 ± 1.95 | 0-8 | **.006** |
| Memory Recall | 0.59 ± 1.30 | 1.74 ± 2.30 | 0-8 | .122 |
| Physically Lost | 0.00 ± 0.00 | 0.83 ± 1.78 | 0-7 | .094 |
| Mentally Lost | 0.55 ± 1.37 | 2.43 ± 2.47 | 0-8 | .070 |
| Past Year (within 12 months) |  |  |  |  |
| Full Blackouts | 0.14 ± 0.47 | 0.43 ± 1.08 | 0-4 | .735 |
| Partial Blackouts | 0.32 ± 0.65 | 1.17 ± 1.90 | 0-5 | .116 |
| Memory Recall | 0.23 ± 0.61 | 1.04 ± 1.80 | 0-5 | .350 |
| Physically Lost | 0.00 ± 0.00 | 0.39 ± 1.20 | 0-5 | .380 |
| Mentally Lost | 0.27 ± 0.77 | 1.39 ± 2.02 | 0-5 | .299 |
| **Psychometric Measures** |  |  |  |  |
| (**DMQR**) Drinking Motives Questionnaire-Revised | | |  |  |
| Social | 11.32 ± 4.52 | 14.82 ± 5.23 | 0-24 | **.023** |
| Enhancement | 7.82 ± 2.36 | 11.50 ± 5.09 | 0-25 | **.014** |
| Coping | 14.41 ± 26.15 | 15.00 ± 23.30 | 0-97 | .171 |
| Conformity | 7.23 ± 3.78 | 7.68 ± 2.82 | 0-19 | .100 |
| (**BIS**) Barrett Impulsivity Scale | | |  |  |
| Motor Impulsiveness | 22.14 ± 3.78 | 21.64 ± 3.79 | 12-32 | .555 |
| Non-planning Impulsiveness | 20.82 ± 4.64 | 21.05 ± 4.21 | 12-30 | .733 |
| Attention Impulsiveness | 15.18 ± 4.48 | 15.86 ± 3.03 | 9-27 | .086 |
| (**STAI**) State and Trait Anxiety Index | | |  |  |
| State Anxiety | 45.00 ± 5.34 | 45.00 ± 4.93 | 35-59 | .680 |
| Trait Anxiety | 44.64 ± 4.72 | 44.32 ± 4.57 | 34-55 | .588 |
| (**VDAQ**) | 37.67 ± 3.16 | 39.38 ± 3.93 | 23-50 | .324 |
| (**COHS**) Creature of Habit Scale | | |  |  |
| Automaticity | 28.78 ± 8.26 | 31.92 ± 5.63 | 16-50 | .357 |
| Routine behaviors | 54.78 ± 6.12 | 51.31 ± 12.55 | 0-69 | .695 |

Values are reported as mean ± standard deviation. Reported *p-*values reflect the results of unpaired two-tailed comparison between groups: Mann-Whitney U non-parametric test for continuous measures or χ^2^ tests for categorical measures. **Boldface** indicates a significant difference between groups at *p*<0.05. VDAQ, Value-Driven Attention Questionnaire

(ARBQ) *Blackouts Total #*: 0, 0; 1, 1; 2, 2; 3, 3; 4, 4-6; 5, 7-11; 6, 12-20; 7, 21-39; 8, 40+

| **Supplemental Table 3. Component Loadings from Categorical Principal Components Analysis of Variables Describing Alcohol Use Behavior and Consequences Under Age 18 among participants who completed the HABIT (N*=*71).** | | | |
| --- | --- | --- | --- |
|  | **Variable** | ***Component 1***  **(42.97%)** | ***Component* 2**  **(39.54%)** |
| **1** | Age of 1^st^ binge episode | **.914** | .221 |
| **2** | Age of 1^st^ drunk experience | **.920** | .213 |
| **3** | Age of 1^st^ drink | **.858** | .031 |
| **4** | Frequency of binge episodes | **.833** | .384 |
| **5** | Total fragmentary blackouts | .329 | **.917** |
| **6** | Prompted recall of lost memories | .307 | **.878** |
| **7** | No memory of actions while drinking night before | .514 | **.794** |
| **8** | Total en block blackouts | -.213 | **.734** |
| **9** | Physically lost while drinking | .492 | **.732** |
| Numbers displayed are loadings after using Varimax rotation. Bolded numbers represent variable weights that contribute to their respective component.  Variables 1, 2, 3 and 4 are from the CAUPQ; 5-9 are from the ARBQ.  Variable 6: Whether lost memories of actions could ever be recalled after prompting  Variable 9: Whether participant has ever found themselves in a place they do not remember going while drinking | | | |

| **Supplemental Table 4. Component Loadings from Categorical Principal Components Analysis of Variables Describing Alcohol Use and Consequences Within the Past Year among participants who completed the HABIT (N*=*71).** | | | |
| --- | --- | --- | --- |
|  | **Variable** | ***Component 1***  **(34.72%)** | ***Component* 2**  **(34.15%)** |
| **1** | Total fragmentary blackouts | **.934** | .263 |
| **2** | No memory of actions while drinking night before | **.931** | .247 |
| **3** | Prompted recall of lost memories | **.858** | .213 |
| **4** | Physically lost while drinking | **.814** | -.075 |
| **5** | Total en bloc blackouts | **.601** | .589 |
| **6** | Times drunk in past 6 months* | .262 | **.840** |
| **7** | Current binge drinking frequency | .367 | **.783** |
| **8** | Percentage of times drunk* | .245 | **.728** |
| **9** | Total drinks on average drinking day | .235 | **.692** |
| **10** | Percentage of days drinking alcohol | -.044 | **.685** |
| **11** | Days drinking per month in past year | -.019 | **.657** |
| Numbers displayed are loadings after using Varimax rotation. Bolded numbers represent variable weights that contribute to their respective component.  *question used to calculate the Townshend and Duka (2009) binge score.  Variables 1, 2, 3, and 4 are from the CAUPQ; 5 and 6 from the CDDR; 7-11 from the ARBQ.  Variable 9: Whether lost memories of actions could ever be recalled after prompting  Variable 10: Whether participant has ever found themselves in a place they do not remember going while drinking | | | |

| **Supplementary Table 5. Component Loadings from Categorical Principal Components Analysis of Variables Describing Alcohol Use and Consequences Within the Past Year among Participants who Completed the RDAB task (N=44).** | | | | | |
| --- | --- | --- | --- | --- | --- |
|  | **Variable** | ***Component 1***  **(41.59%)** | | ***Component* 2**  **(34.15%)** | |
| **1** | Times drunk in past 6 months* | | **.912** | | .194 |
| **2** | Total drinks on average drinking day | | **.833** | | .272 |
| **3** | Current binge drinking frequency | | **.780** | | .491 |
| **4** | Percentage of times drunk* | | **.759** | | .195 |
| **5** | Total en bloc blackouts | | **.752** | | .480 |
| **6** | Days drinking per month in past year | | **.735** | | .025 |
| **7** | Percentage of days drinking alcohol | | **.700** | | .059 |
| **8** | Total fragmentary blackouts | | .306 | | **.930** |
| **9** | No memory of actions while drinking night before | | .301 | | **.920** |
| **10** | Prompted recall of lost memories | | .289 | | **.859** |
| **11** | Physically lost while drinking | | -.051 | | **.826** |
| Numbers displayed are loadings after using Varimax rotation. Bolded numbers represent variable weights that contribute to their respective component.  *question used to calculate the Townshend and Duka (2009) binge score.  Variables 1, 2, 3, and 4 are from the CAUPQ; 5 and 6 from the CDDR; 7-11 from the ARBQ.  Variable 10: Whether lost memories of actions could ever be recalled after prompting  Variable 10: Whether participant has ever found themselves in a place they do not remember going while drinking | | | | | |

| **Supplemental Table 6. Component Loadings from Categorical Principal Components Analysis of Variables Describing Alcohol Use Behavior and Consequences Under Age 18 among Participants who Completed the RDAB Task (N*=*44).** | | | |
| --- | --- | --- | --- |
|  | **Variable** | ***Component 1***  **(47.95%)** | ***Component* 2**  **(32.81%)** |
| **1** | Age of 1^st^ drunk experience | **.939** | .141 |
| **2** | Age of 1^st^ binge episode | **.927** | .182 |
| **3** | Age of 1^st^ drink | **.873** | -.041 |
| **4** | Frequency of binge episodes | **.850** | .262 |
| **5** | No memory of actions while drinking night before | **.625** | .558 |
| **6** | Total fragmentary blackouts | .188 | **.939** |
| **7** | Total en block blackouts | -.104 | **.847** |
| **8** | Physically lost while drinking | .417 | **.770** |
| Numbers displayed are loadings after using Varimax rotation. Bolded numbers represent variable weights that contribute to their respective component.  Variables 1, 2, 3 and 4 are from the CAUPQ; 5-8 are from the ARBQ.  Variable 8: Whether participant has ever found themselves in a place they do not remember going while drinking | | | |

|  | **Supplemental Table 7**. Effect of reward on performance during reward conditioning (N=44). | | | | | | |
| --- | --- | --- | --- | --- | --- | --- | --- |
|  |  | **Adolescent Alcohol Use** | | | | | |
|  |  | **Light** | | | **Heavy** | | |
| **Block #** | **Reward Value** | **RT** | **Accuracy** | **Inverse Efficiency** | **RT** | **Accuracy** | **Inverse Efficiency** |
| **B1** | Rewarded | 614 (50) | 0.87 (0.10) | 720 (167) | 584 (34) | 0.89 (0.08) | 661 (83) |
|  | Unrewarded | 613 (49) | 0.81 (0.13) | 779 (191) | 585 (32) | 0.83 (0.12) | 725 (152) |
| **B2** | Rewarded | 584 (43) | 0.88 (0.10) | 676 (142) | 583 (36) | 0.89 (0.08) | 659 (89) |
|  | Unrewarded | 589 (43) | 0.81 (0.13) | 745 (162) | 587 (35) | 0.83 (0.12) | 730 (158) |

SD = Standard Deviation. All means and SD report reaction times in milliseconds.

| **Supplemental Table 8. Hierarchical Linear Multiple Regression Analysis of adolescent and past year alcohol use on reward conditioning during the RDAB task (N=44).** | | | | | |
| --- | --- | --- | --- | --- | --- |
| **Variable** | **B** | **SE** | ***β*** | ***p-value*** | **95% CI** |
| ***Step 1-* covariates** |  |  |  |  |  |
| Recency of alcohol consumption | .001 | .002 | .119 | .382 | [-.009 .005] |
| ***Step 2 -* predictors** |  |  |  |  |  |
| Heavy adolescent alcohol use | -.030 | .924 | -.015 | .977 | [-2.005, 1.491] |
| Past year alcohol use | .018 | .015 | .236 | .245 | [-.012, .047] |
| Adolescent X Past year alcohol use | -.003 | .026 | -.041 | .906 | [-.043, .057] |
| *Note:* Heavy adolescent alcohol is a binary variable; 0, light use; 1, heavy use.  B, unstandardized coefficient; *β****,*** standardized coefficient; *SE,* standard error. *p-value* based on 1000 bootstrap samples. **p*<0.05 | | | | | |

| **Supplemental Table 9**. Performance during the attention cueing paradigm for rewarded trials (N=44). | | | | | | | |
| --- | --- | --- | --- | --- | --- | --- | --- |
|  |  | **Adolescent Alcohol Use** | | | | | |
|  |  | **Light** | | | **Heavy** | | |
| **Cueing Type** | **Run #** | **RT** | **Accuracy** | **Inverse Efficiency** | **RT** | **Accuracy** | **Inverse Efficiency** |
| **Valid** | **R1** | 519 (59) | 0.94 (8.7) | 562 (121) | 510 (36) | 0.97 (3.8) | 528 (47) |
|  | **R2** | 509 (45) | 0.95 (6.5) | 541 (94) | 502 (41) | 0.96 (5.7) | 528 (73) |
|  | **R3** | 510 (44) | 0.95 6.8) | 542 (100) | 507 (38) | 0.95 (4.6) | 534 (65) |
| **Neutral** | **R1** | 520 (58) | 0.94 (8.7) | 564 (120) | 513 (36) | 0.97 (4.2) | 532 (50) |
|  | **R2** | 512 (45) | 0.95 (7.0) | 548 (101) | 506 (41) | 0.95 (6.5) | 536 (80) |
|  | **R3** | 509 (44) | 0.95 (7.3) | 544 (107) | 506 (37) | 0.96 (4.7) | 532 (66) |
| **Invalid** | **R1** | 519 (59) | 0.94 (8.9) | 565 (124) | 511 (37) | 0.97 (4.0) | 530 (50) |
|  | **R2** | 511 (45) | 0.95 (7.1) | 547 (105) | 506 (41) | 0.95 (6.0) | 535 (76) |
|  | **R3** | 509 (44) | 0.95 (6.7) | 541 (97) | 507 (38) | 0.95 (4.7) | 535 (66) |

SD = Standard Deviation. All means and SD report reaction times in milliseconds.

| **Supplemental Table 10**. Performance during the attention cueing paradigm for unrewarded trials (N=44). | | | | | | | |
| --- | --- | --- | --- | --- | --- | --- | --- |
|  |  | **Adolescent Alcohol Use** | | | | | |
|  |  | **Light** | | | **Heavy** | | |
| **Cueing Type** | **Run #** | **RT** | **Accuracy** | **Inverse Efficiency** | **RT** | **Accuracy** | **Inverse Efficiency** |
| **Valid** | **R1** | 519 (59) | 0.94 (0.09) | 563 (121) | 511 (36) | 0.97 (0.04) | 530 (49) |
|  | **R2** | 510 (44) | 0.95 (0.07) | 542 (96) | 504 (41) | 0.96 (0.06) | 531 (74) |
|  | **R3** | 509 (43) | 0.95 (0.07) | 541 (100) | 505 (37) | 0.96 (0.05) | 530 (63) |
| **Neutral** | **R1** | 520 (58) | 0.94 (0.09) | 564 (120) | 513 (36) | 0.97 (0.04) | 532 (50) |
|  | **R2** | 512 (45) | 0.95 (0.07) | 548 (101) | 506 (41) | 0.95 (0.07) | 536 (80) |
|  | **R3** | 509 (44) | 0.95 (0.07) | 544 (107) | 506 (37) | 0.96 (0.05) | 532 (66) |
| **Invalid** | **R1** | 521 (59) | 0.93 (0.10) | 573 (134) | 514 (37) | 0.96 (0.05) | 537 (56) |
|  | **R2** | 509 (44) | 0.95 (0.07) | 542 (92) | 503 (42) | 0.96 (0.06) | 530 (75) |
|  | **R3** | 508 (44) | 0.95 (0.07) | 539 (96) | 505 (37) | 0.96 (0.05) | 531 (64) |

SD = Standard Deviation. All means and SD report reaction times in milliseconds.
